# Validation of an Assessment Scale for a Low-Tech Laparoscopic Appendectomy Simulation and Its Relevance for Formative Self-Assessment

**DOI:** 10.64898/2026.07.20.26358477

**Authors:** Hugues Thierry Tumameu Kouam, Louise Renoult, Maëlig Poitevin, Lucie Jourdin, Clara Hervé, Pierre Meignan, Guillaume Podevin, Françoise Schmitt

## Abstract

**Introduction:** Laparoscopic appendectomy is an ideal procedure for acquiring laparoscopic skills through simulation. Nevertheless, technical training is time consuming for surgical trainers to provide constructive feedback, but this could be improved by the development of validated tools that enable appropriate formative self-assessment. For this reason, we developed a structured assessment scale for a laparoscopic appendectomy exercise using a low-fidelity simulator. The objective of this study was to validate the scale for use in formative self-assessment.

**Methods:** During laparoscopic simulation sessions in 2025–2026, participants with varying levels of experience performed a standardized laparoscopic appendectomy (LAP) exercise on a low-fidelity simulator. Performance was assessed through formative self- and external assessment using a specific scale derived from the OSATS (Objective Structured Assessment of Technical Skills) score. Content and construct validity, internal consistency, reproducibility, and reliability in both hetero- and self-assessment were analyzed.

**Results:** Thirty-two participants were included in the validation study of the LAP scale, including 7 medical students, 17 residents in pediatric, visceral, urological, and gynecological surgery, and 8 practicing surgeons. The content of the scale was deemed relevant by 80% of the users. It demonstrated excellent construct validity, with scores increasing according to level of experience: 9.9 ± 0.7 among students, 12.7 ± 3.3 among junior residents, 16.6 ± 3.3 among experienced residents, and 18.8 ± 0.9 among practicing surgeons (p < 0.0001). Reproducibility and internal consistency were significant, while inter- and intra-rater reliability were excellent (correlation coefficients r = 0.90 and 0.91; p < 0.0001), as was the correlation between external and self-assessment (r = 0.81; p < 0.0001). Self-assessment was more reliable among experienced learners than among novices.

**Conclusion:** This standardized LAP scale is validated for both external and self-assessment, the latter requiring prior training to be reliable and formative.

## 1. Introduction

Acute appendicitis is one of the most common surgical emergencies of digestive surgery in both children and adults [1], and its treatment involves an appendectomy [2]. When equipment and expertise are available, laparoscopic appendectomy (LAP) is considered the preferred approach due to its advantages in terms of morbidity, length of hospital stay and postoperative pain [3, 4]. It is therefore one of the first and most frequently performed procedures during residency training [5].

However, so-called “minimally invasive” surgical techniques require extensive training to reach an expert level, which is increasingly achieved through simulation-based training that complements hands-on experience in the operating room [6]. This teaching method is recognized as particularly effective for acquiring and retaining technical skills, but it requires specialized equipment and a significant investment of instructors’ time. To overcome these challenges, various strategies can be implemented, such as the development of low-cost “do-it-yourself” training models [7, 8], which allow for the repetition of technical skills at a lower cost compared to commercial models, and the development of peer-to-peer or self-directed training sessions [9, 10]. The effectiveness of these training sessions appears to be underpinned by prior learning and regular reassessment sessions with an instructor, as well as by the availability of validated formative assessment tools for both external and self-assessment [11].

We have developed an evaluation tool for a laparoscopic appendectomy exercise on a low-fidelity simulator, based on the OSATS (Objective Structured Assessment of Technical Skills) score [12]. This educational simulator allows for training in sequencing various technical skills required during an appendectomy, including the identification and bimanual mobilization of structures, dissection of the mesoappendix with scissors, hemostasis control (here using clips), ligation of the appendiceal base using an endoloop or internal knot, and proper resection of the appendix.

The aim of this study was to validate this LAP assessment scale as a formative self-assessment tool, after testing its criteria for content validity, discriminant validity, reliability, and reproducibility.

## 2. METHODS

### 2.1. Ethical Considerations

This study, which did not involve health data, was conducted after obtaining approval from the Ethics Committee of the Angers University Hospital (n°2025-050, March 12, 2025). The study was registered with the French Data Protection Authority (CNIL) under reference methodology MR004. As this was an educational study, it was not registered in a clinical trial registry. Large language models (LLM) were used to optimize the literature review prior to this study and to improve its English writing. After using it, the authors reviewed and edited the content as needed and take full responsibility for the content of the published article.

### 2.2. Participants

All participants in this prospective cohort study were included on a voluntary basis, following oral and written information and the collection of written consent and authorization to record videos.

The study took place from December 2024 to March 2026 during laparoscopy training sessions at the health simulation centers of the Angers and Tours University Hospitals, All’Sims *(Angers Loire Learning SIMulation en Santé)* and MEDISIM. Participants included medical students, junior surgical residents (first two years of residency) and senior surgical residents (^3rd^and^4th^years), as well as attending surgeons. The minimum number of participants required to assess the discriminatory criterion between two groups with different levels of expertise was calculated *a priori* using the online software BiostaTGV (Jussieu, France [biostatgv.sentiweb.fr]) and was set at 7 participants per group, for a mean difference of 3 points on the LAP scale, with a Type I error rate of 5% and a power of 0.8.

### 2.3. Simulated Laparoscopic Appendectomy Technique and Assessment Scale

Trainees had access to standard laparoscopic instruments, including atraumatic forceps, scissors, clips, and a slipknot simulating an Endoloop™ ligature; all of these were used on an FLS laparotrainer (Fundamental Skills in Laparoscopy, Medicalem S.A.S. St Cyr l’Ecole, France), with a low-cost appendectomy model.

The surgical procedure proposed in this exercise consisted, in order, of:

a. dissection of the mesoappendix with scissors to isolate the artery
b. ligation of the appendicular artery with clips, followed by its transection between these clips
c. placement of a slipknot at the base of the appendix
d. transection of the appendix

Some exercises were filmed anonymously, with the participant’s consent, to allow for iterative filling of the LAP scale remotely.

At the end of the exercise, each participant completed as self-assessment (SA) the LAP scale, a 20-point evaluation form for formative purposes (Table 1), adapted from the OSATS (Objectives Structured Assessment of Technical Skills) score [12]. The same evaluation form was completed at the end of the exercise by one or two independent evaluators as part of an external assessment (EA). The exercise was considered successful when the final score was greater than or equal to 12.5 out of 20 points, corresponding to the minimum competency threshold of +0.5 points for the technical items (A–E) assessing surgical efficacy and safety, and +0 points for the more general items assessing surgical knowledge and ergonomics (items F–J).

**Table 1:**
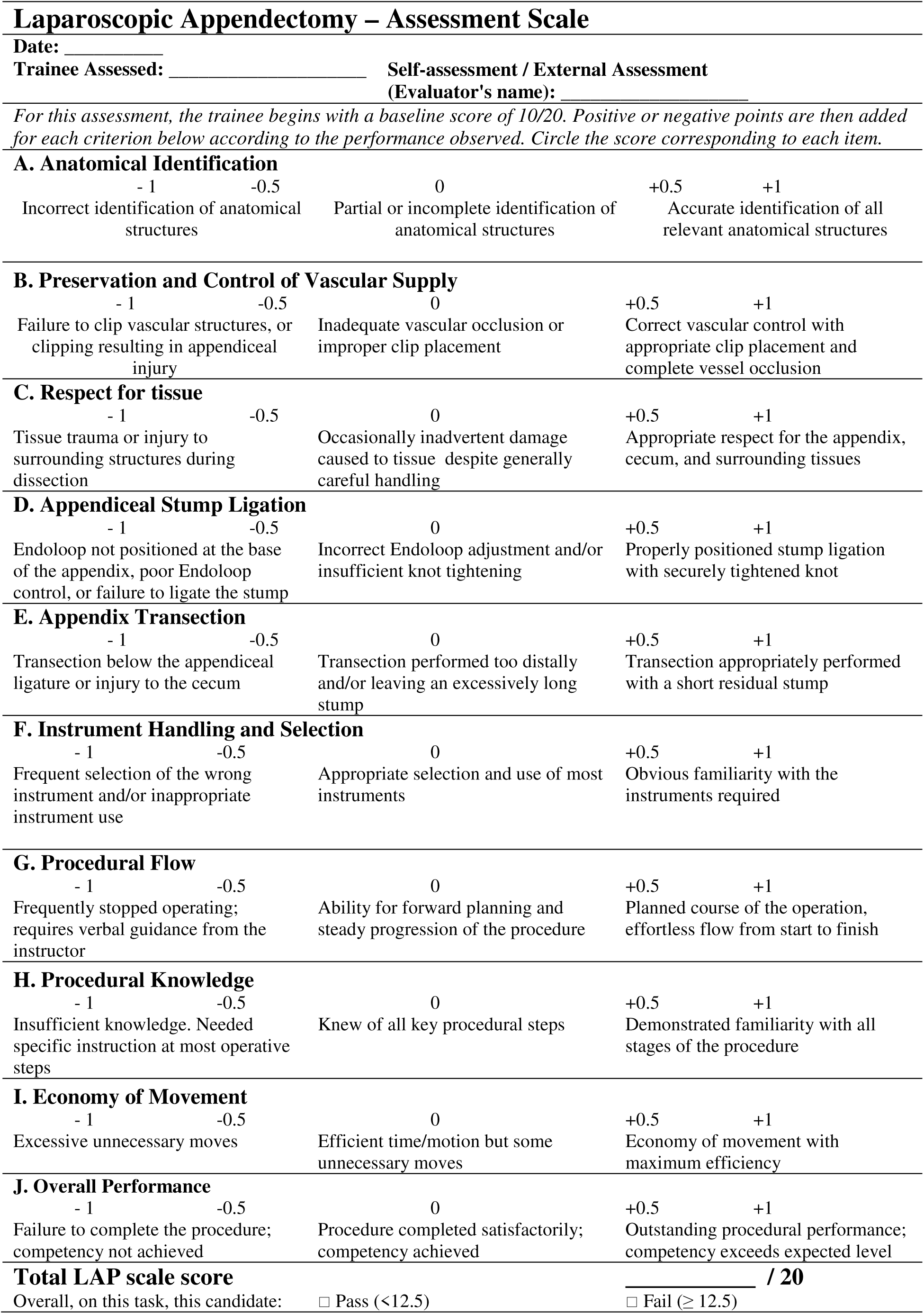
Laparoscopic Appendectomy assessment scale.

Participants and evaluators were also asked to complete a digital questionnaire (Microsoft Forms) regarding their satisfaction with the assessment tool.

### 2.4. Validation of the LAP Assessment scale

The evaluation tool was validated using the following six parameters, based on participants’ first attempt at the exercise:

- Content validity was tested by having the various users of the LAP scale complete an online satisfaction questionnaire (Microsoft Forms).
- Construct validity was assessed by comparing the median scores obtained by groups of novices (medical students), junior surgical residents (years 1 and 2 of training), and senior surgical residents (years 3 and 4 of training), as well as by experts (attending surgeons);
- Reproducibility was tested by analyzing the median scores of first-year residents, who performed the LAP exercise under the same conditions, after dividing them into two comparison groups matched on their prior surgical experience, as estimated by the number of weeks of surgical rotations during medical school.
- Inter-rater reliability was assessed by correlating the scores of two independent external evaluators;
- Intra-rater (or test-retest) reliability was assessed through a blinded analysis of anonymized videos taken during the exercises by three expert surgeons, repeated more than one month apart.
- The internal consistency of the multimodal LAP scale was calculated using Cronbach’s α.
- Finally, the reliability of the scale when used for self-assessment was tested by analyzing the correlation between self-assessment and external assessment scores.

### 2.5. Biostatistical Analysis

Statistical analysis was performed using the Rplusplus (Zebrys, France) and GraphPad Prism 8.0.2 for Windows software packages. Quantitative variables were expressed as means and standard deviations or medians and interquartile ranges, depending on the nature of the variable, and qualitative data were expressed as frequencies or percentages. Comparative statistical analyses were performed at a 5% significance level (α) using Student’s t-test or the Mann-Whitney U-test for binary quantitative data, depending on whether the data followed a normal distribution as confirmed by the Shapiro-Wilk test, and the Kruskal-Wallis test for comparisons involving three or more groups, combined with Dunn’s multiple comparison test. Qualitative data were compared using the chi-square or Fisher’s exact tests, and correlation coefficients were calculated using Spearman’s or Pearson’s correlation tests depending on the nature of the variable; a correlation coefficient r > 0.7 was considered significant. Cronbach’s alpha was calculated using SPSS software for Windows.

## 3. Results

### 3.1. Description of participants

Assessment data from 32 participants were analyzed. These participants were divided into 7 novices, 17 surgical residents (including 10 junior and 7 senior surgical residents), and 8 attending surgeons. Four surgical specialties were represented: gastrointestinal surgery (24%), pediatric surgery (20%), urology (24%), and gynecology (32%).

### 3.2. Validation criteria for the assessment scale

The relevance of the LAP scale for evaluating a low-tech simulation of an appendectomy was assessed by 20 observers, with an average score of 8.3 ± 1.2 out of 10 points; 80% of them considered it very suitable for evaluating this exercise. Only 60% of them considered it very useful for distinguishing a student’s level of experience, with an average score of 7.6 ± 1.8 out of 10 points. Nevertheless, analysis of the results obtained by the four groups with different levels of experience revealed a gradual and significant increase in the median score out of 20 points, rising from 9.8 [9.4 – 10.4] among novices to 19.0 [18.3 – 19.3] among attending surgeons (Figure 1a), and success rates ranging from 0% among novices to 100% among attending surgeons, with a gradual increase among junior residents (50%) and senior residents (85%).

**Figure 1:**
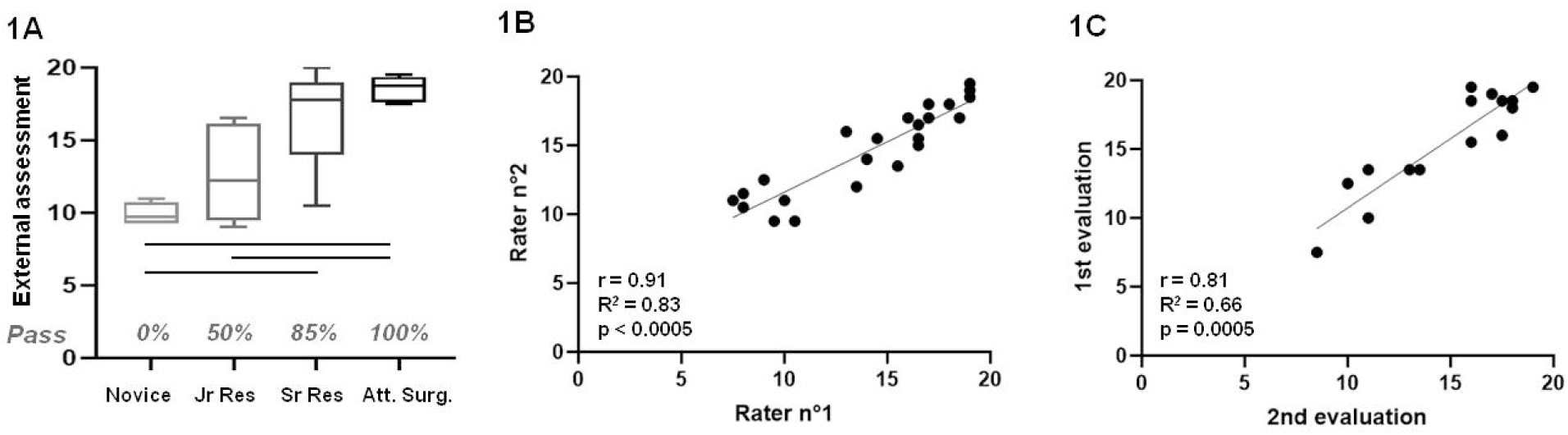
Validity of the assessment scale for simulated laparoscopic appendectomy. Figure 1A: Construct validity. Jr Res = junior surgical residents; Sr Res = senior surgical residents; Att. Surg = attending surgeons. Gray italicized numbers: exercise pass rate (“Pass”). Figure 1B: Inter-rater reliability. Figure 1C: Intra-rater reliability. r = correlation coefficient; R^²^= coefficient of determination.

During laparoscopic training sessions, which included participants with varying levels of expertise, the exercise was scored independently by two external evaluators on 23 occasions. The correlation coefficient between these evaluations was r = 0.91 [0.78–0.96], p < 0.0005 (Figure 1B), indicating good inter-observer reliability.

To analyze intra-observer (or test-retest) reliability, anonymized videos of 5 participants were recorded and scored in a blinded manner on two separate occasions, more than two weeks apart, by three independent experts. The correlation coefficient obtained was r = 0.81 [0.49–0.94], p = 0.0005 (Figure 1C).

Reproducibility was tested by comparing the median scores of two groups of six first-year residents, matched for their prior surgical experience, as estimated by the number of weeks of clinical rotations in surgical departments completed during their medical studies. The two groups were equivalent in terms of age, sex, distribution of specialties, and prior surgical experience (Table 2). During their first simulated laparoscopic appendectomy, the median scores were equivalent in both peer assessment (13.8 vs. 14.6, p = 0.68) and self-assessment (12.0 vs. 11.3, p = 0.15).

**Table 2:** Comparative Analysis of Two Groups of First-Year Residents.

|  | **Group 1 (n = 6)** | **Group 2 (n = 6)** | **p-value** |
| --- | --- | --- | --- |
| Age | 24 [24 – 25.3] | 24 [23.8 - 25] | 1 |
| Male (n (%)) | 4 (66.7%) | 1 (16.7%) | 0.24 |
| Right-handed (n (%)) | 6 (100%) | 5 (83.3%) | 1 |
| Graduating class (2024/2025) | 3/3 | 3/3 | 1 |
| Median number of surgical rotations | 4.5 [2.5 – 5.3] | 3 [3 - 4.3] | 0.42 |
| Median durations of surgical rotations (weeks) | 21.5 [16 – 49] | 22 [13.8 – 44.5] | 1 |
| Specialty (n (%)) |  |  | 0.5 |
| Gynecology | 1 (16.7%) | 2 (33.3%) |  |
| Pediatric surgery | 0 (0%) | 1 (16.7%) |  |
| Urology | 2 (33.3%) | 2 (33.3%) |  |
| Gastrointestinal Surgery | 3 (50.0%) | 1 (16.7%) |  |
| Median score on external assessment | 13.8 [9.8 – 16.1] | 14.6 [12.3– 15.4] | 0.68 |
| Pass rate in peer assessment | 66.7% | 83.3% | 1 |
| Median score on self-assessment | 12 [11.8 – 13] | 11.3 [9.4– 12.6] | 0.15 |
| Self-assessment success rate | 33.3% | 16.7% | 1 |

Finally, the Cronbach’s alpha coefficient was α = 0.866, indicating very good internal consistency.

### 3.3 Reliability of the formative self-assessment scale

As with external assessment, the analysis of surgical-level subgroups showed a gradual and significant increase in the median scores on the LAP-scale (p < 0.0001), rising from 9.5 [8.3 – 11.0] among novices to 17.5 [16.9 – 18.1] for experts, with improving success rates (Figure 2A). The internal consistency of the self-assessment scale was equivalent to that of the inter-rater assessment (α = 0.847). Across the entire cohort, the correlation coefficient between self-assessment and external assessment was r = 0.83 [0.65 – 0.92], p < 0.0001 (Figure 2B). A subgroup analysis (Figure 2C) showed that this correlation improved with increasing learner expertise; it was not significant among novices (r = −0.34, p = 0.45) and junior residents (r = 0.21 [-0.32 – 0.82], p = 0.55), but very high among senior residents (r = 0.94 [0.62 – 0.99], p = 0.002).

**Figure 2:**
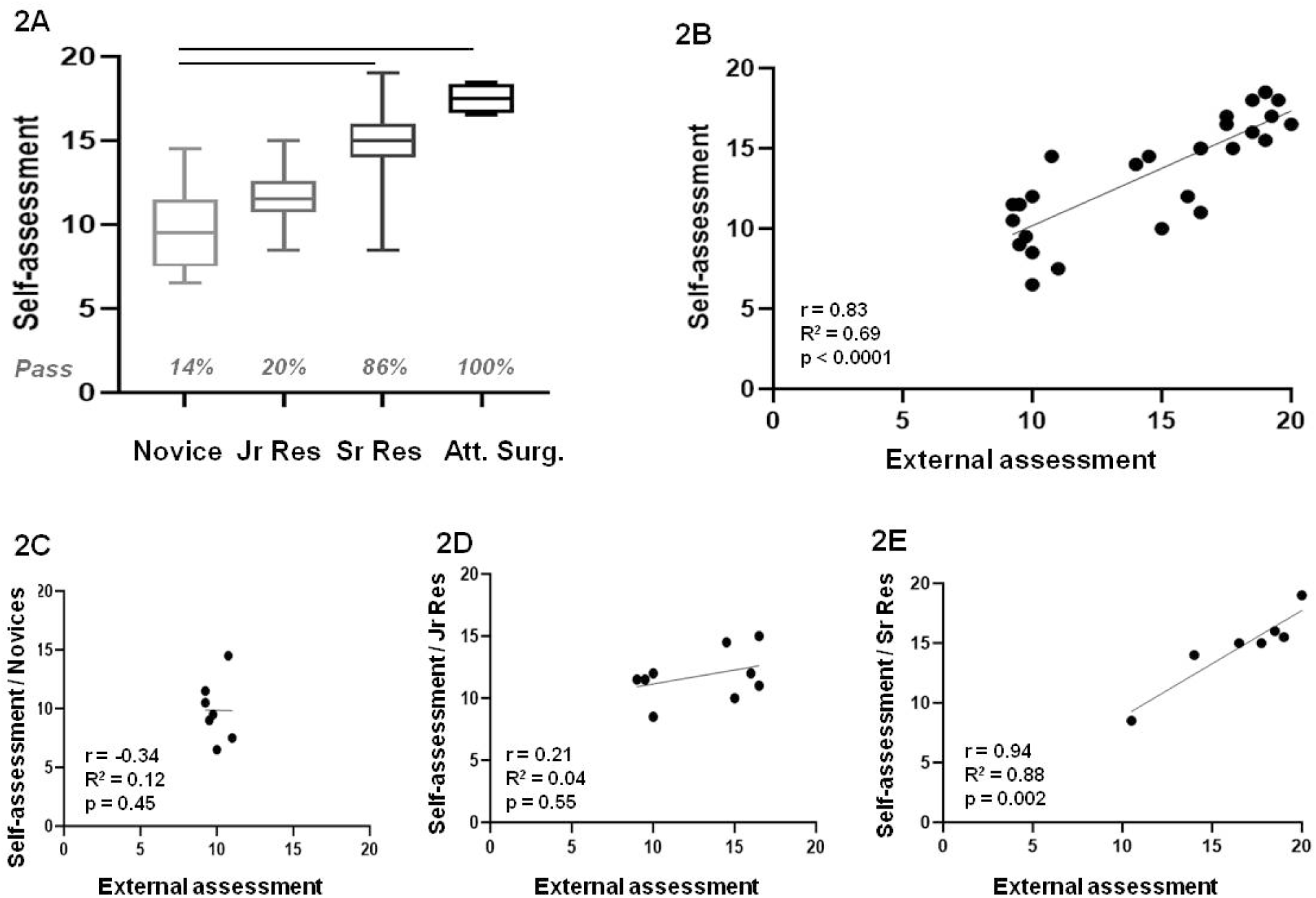
Validation of the simulated laparoscopic appendectomy scale for self-assessment. Figure 2A: Construct validity of the self-assessment scale. Gray numbers: success rate on the exercise (“Pass”). Figure 2B: Overall correlation between self-assessment and external assessment. r = correlation coefficient; R^²^= coefficient of determination. Figures 2C, 2D, 2E: Correlation between self-assessment and external assessment in subgroups of novices, junior residents (Jr Res), and senior residents (Sr Res).

## 4. Discussion

This study thus validates a specific laparoscopic appendectomy skills assessment scale for both external and self-assessment using a low-tech simulation model. Indeed, the reproducibility and test-retest reliability showed very good scores in external assessment. Internal consistency, content and construct validities were also validated, both in expert evaluations and in self-assessments. Inter-observer reliability among experts showed a high and significant correlation coefficient, but proved to be slightly less robust in self-assessments, particularly among the least experienced participants.

Appendectomy is one of the laparoscopic procedures that surgical residents can perform most quickly [13]. It is also a simple procedure that lends itself well to simulation training and allows for the acquisition of various basic laparoscopic technical skills, such as bimanual coordination, dissection with scissors, and the placement of clips or slip knots. For this reason, many teams have recently focused on developing digital or low-tech models using a variety of materials, ranging from inverted gloves [14, 15] to 3D silicone models [16, 17]. Most of them have incorporated this exercise into their educational curriculum and developed associated assessment tools. A recent meta-analysis [18] lists 12 tools used to evaluate laparoscopic appendectomy, including 6 adapted for high-fidelity digital simulators, 3 generic scales (GOALS, modified GOALS, and OSATS) and 3 specific scales; among the latter, the LARS scale [19] is validated for real surgical appendectomies, while the team led by Adrales *et al.* [20] uses a generic 4-item scale, and Skjold’s scale [21] focuses on the varying degrees of autonomy of the trainee surgeon during the procedure. More recently, three new scales have been described: the LapPASS [22] incorporates an assessment of the appendicular base ligation, a tool specific to the LapAppendectomy4All learning model [17] uses a generic surgical competency score, and the APPY-VOP composite assessment scale has demonstrated good construct validity and concurrent validity with the mOSATS scale [15]. The latter is likely the assessment tool most similar to the one proposed in this study, as it incorporates a checklist and generic competency items. Nevertheless, their main difference lies in the scoring of technical items that occur sequentially during the appendectomy procedure: while the APPY-VOP tool uses a binary checklist, our scale immediately associates the performance of each step of the procedure with an assessment of the quality of that step’s execution, thereby aligning more closely with generic assessment tools, which are generally described as more effective for scoring technical competence [23].

In this study, the assessment scale for the laparoscopic appendectomy exercise meets all the validity criteria recommended by the MERSQI score [24], with the exception of concurrent validity. This omission is due to the lack of a relevant external reference: to our knowledge, no published assessment tool is specifically designed for this exercise while strictly adhering to the expected procedure in our model, which prevents the identification of a true gold standard. Thus, the APPY-VOP tool [15] exhibits notable discrepancies regarding the steps of dissection and vascular ligation, making any direct comparison inappropriate without prior adaptation. Furthermore, the use of a validated global scale could have been considered from a purely methodological standpoint. However, this option was not selected, as it would have made the educational program more cumbersome without allowing for a relevant assessment of concurrent validity for items specifically related to the procedure.

Based directly on the OSATS score, the design of the LAP-scale focused on incorporating 7 of the 8 general surgical competency items, omitting the one regarding the use of an assistant, which was unnecessary for this exercise, combining the items on knowledge and handling of instruments into a single item, and developing five procedure-specific items, thereby enabling a detailed assessment of performance at each critical stage of the procedure. Based on our faculty’s grading principles, the items were scored positively or negatively relative to the mean, rather than on a scale of 1 to 5 as in the original score, with the aim of limiting the ceiling effect and better aligning with competency-based surgical education and the concept of entrustable professional activities (EPAs) [25], which focus on the progressive nature of learning and the expected level of competency.

While several scoring tools for a laparoscopic appendectomy exercise have been developed, none has yet been tested and validated for use in self-assessment or peer assessment, even though this has been suggested by some authors [15]. The LAP-scale demonstrated good construct validity in self-assessment, as well as a strong correlation between self-assessment and external assessment. Nevertheless, a clear improvement in the reliability of self-assessment was observed with the learner’s level of expertise: novices showed no satisfactory correlation with the expert assessor.

Self-assessment appears to be influenced by numerous factors, including both extrinsic factors - such as the complexity of the technical task [26] or the type of assessment scale [27] - and intrinsic factors - such as self-confidence [28] and personal experience with the task [29, 30], which helps optimize alignment with the expected “expert” level through a better understanding and assessment of that level. No particular tendency toward systematic over- or underestimation by the most novice participants was found here, which seems to contradict the existence of a significant Dunning-Kruger effect - an effect that has nevertheless been widely described in surgical education [31] and which may limit the possibility of conducting a reliable self-assessment [32, 33]. These results may therefore support the possibility of effectively using self-assessment guided by standardized and validated tools, provided that trainees first learn how to use them properly. This could be achieved through videos narrated by experts, practice sessions conducted and/or observed among peers of different technical levels combined with feedback from an instructor, and a debriefing with an instructor regarding the self-assessment performed by the most novice participants in order to improve the accuracy of their judgment by developing their metacognitive abilities [10].

The purpose of this study was to validate the LAP-scale for virtually all items listed by Messick [34], with the exception of concurrent validity, due to the absence of an appropriate gold standard. This allows us to propose a competency assessment tool classified at Level 2 on the Kirkpatrick scale [35], validated by a rigorous methodology modeled on the MERSQI criteria [24], and developed within a suitable environment by a team experienced in surgical education research. Its main weakness lies in the small sample sizes of the expertise subgroups, which could lead to potential selection bias, despite *a priori* calculation of the number of participants needed to ensure satisfactory statistical power.

## 5. Conclusion

The results of this study therefore allow us to propose a specific and validated tool for evaluating a standardized low-tech laparoscopic appendectomy exercise. The description of the various technical criteria should allow it to be adapted to most synthetic appendectomy simulators currently described in the literature. Its use for formative self-assessment has also been validated, provided that the most novice learners receive prior training in its use.

## Supporting information

Supplemental Table 1 - French version

## Data Availability

All data produced in the present study are available upon reasonable request to the authors

