## Supplemental Table 1 - French version for "Validation of an Assessment Scale for a Low-Tech Laparoscopic Appendectomy Simulation and Its Relevance for Formative Self-Assessment"

### Appendicectomie laparoscopique
### Score OSATS

**Date :** **Etudiant évalué :**

Autoévaluation / Hétéroévaluation (nom de l'évaluateur) :

**Score OSATS** (Objective Structural Assessment of Technical Skills): **_______ / 20**

*Dans cette évaluation, l'apprenant commence avec une notre de 10/20.*
*S'ajouteront alors pour chaque item ci-dessous les points positifs et négatifs selon les actions réalisées.*
*Entourer pour chaque item ci-dessous le chiffre correspondant.*

**A] Repérage anatomique :**

| -1 | -0,5 | 0 | +0,5 | +1 |
|---|---|---|---|---|
| Mauvaise identification des structures anatomiques | | Identification partielle ou imparfaite des structures | | Identification correcte de toutes les structures |

**B] Respect/clampage de la vascularisation :**

| -1 | -0,5 | 0 | +0,5 | +1 |
|---|---|---|---|---|
| Pas de clampage, ou clampage avec atteinte de l'appendice | | Clampage perméable, ou mauvais positionnement des clips | | Clampage correctement effectué (bonne localisation et imperméabilité) |

**C] Respect des tissus :**

| -1 | -0,5 | 0 | +0,5 | +1 |
|---|---|---|---|---|
| Fragilisation ou lésion des tissus et structures au cours de la dissection | | Quelques erreurs fragilisant les tissus malgré une préhension prudente et appropriée | | Respect de l'appendice, du caecum et des tissus environnants |

**D] Ligature de l'appendice :**

| -1 | -0,5 | 0 | +0,5 | +1 |
|---|---|---|---|---|
| Endoloop non placé à la base de l'appendice / Pas de tenue de l'endoloop ou absence de ligature du moignon | | Mauvais ajustement de l'endoloop / nœud pas assez serré | | Ligature du moignon bien positionnée et nœud correctement serré |

**E] Section de l'appendice :**

| -1 | -0,5 | 0 | +0,5 | +1 |
|---|---|---|---|---|
| Section en-dessous de la ligature du moignon appendiculaire ou lésion du caecum | | Section trop basse et/ou moignon trop long | | Section bien réalisée , avec petit moignon |

FACULTÉ DE SANTÉ
UNIVERSITÉ D'ANGERS

AllSims
Angers Loire Learning Simulation Santé

**F] Utilisation des instruments :**

| -1 | -0,5 | 0 | +0,5 | +1 |
|---|---|---|---|---|
| Choix fréquent du mauvais instrument et/ou utilisation d'instruments inadaptés au geste réalisé | | Choix correct de la plupart des instruments et utilisation adaptée de ceux-ci | | Familiarité évidente avec le nom et l'utilisation de tous les instruments |

**G] Déroulement de la procédure :**

| -1 | -0,5 | 0 | +0,5 | +1 |
|---|---|---|---|---|
| Procédure arrêtée fréquemment, impression de mauvaise connaissance de l'étape suivante, nécessité d'une aide orale du formateur | | Progression de la procédure raisonnable avec anticipation et planification des étapes | | Procédure parfaitement planifiée et efficace du début à la fin |

**H] Connaissance de la procédure :**

| -1 | -0,5 | 0 | +0,5 | +1 |
|---|---|---|---|---|
| Connaissance insuffisante semblant non sûre, présence d'hésitations | | Connaissance de toutes les étapes importantes de la procédure | | Aisance et habitude de l'ensemble des étapes de la procédure |

**I] Gestuelle :**

| -1 | -0,5 | 0 | +0,5 | +1 |
|---|---|---|---|---|
| Beaucoup de gestes inutiles | | Gestuelle efficace mais encore parasitée par quelques mouvements inutiles | | Economie nette de mouvements avec résultat optimal |

**J] Performance globale :**

| -1 | -0,5 | 0 | +0,5 | +1 |
|---|---|---|---|---|
| Echec de réalisation de la procédure, compétence non acquise | | Réalisation correcte de la procédure, compétence acquise | | Réalisation parfaite de la procédure, compétence supérieure |

Exercice validé(≥12,5/20)? Oui / Non
